# A large-scale machine learning study of sociodemographic factors contributing to COVID-19 severity

**DOI:** 10.1101/2023.01.27.23285043

**Authors:** Marko Tumbas, Sofija Markovic, Igor Salom, Marko Djordjevic

## Abstract

Understanding sociodemographic factors behind COVID-19 severity relates to significant methodological difficulties, such as differences in testing policies and epidemics phase, as well as a large number of predictors that can potentially contribute to severity. To account for these difficulties, we assemble 115 predictors for more than 3000 US counties and employ a well-defined COVID-19 severity measure derived from epidemiological dynamics modeling. We then use a number of advanced feature selection techniques from machine learning to determine which of these predictors significantly impact the disease severity. We obtain a surprisingly simple result, where only two variables are clearly and robustly selected - population density and proportion of African Americans. Possible causes behind this result are discussed. We argue that the approach may be useful whenever significant determinants of disease progression over diverse geographic regions should be selected from a large number of potentially important factors.

## 1 Introduction

More than two years into the COVID-19 pandemic, there are still many open questions regarding the spread and severity of SARS-CoV-2. Not only can we not explain, on an individual basis, who will experience severe illness or no symptoms at all, but we often lack this predictive power even on the larger scale of entire regions, where personal traits and individual genetical predispositions are averaged out. Different countries or regions within a country experience diverse numbers of new cases and fatalities, with patterns that are difficult to anticipate. On the other hand, the potential benefits of the ability to understand and foresee the regional COVID-19 behavior are clear: it would assist governments in appropriately allocating resources, help sustain economic activities, and allow to correctly and timely estimate risks and necessary measures - thus saving human lives and reducing the overall epidemic impact.

Naively, in the present era of abundant and widely available data, one could expect that most of these questions could be settled down by systematically comparing the COVID-19 numbers with various demographic and environmental parameters. However, while much progress in this direction has been made (e.g., (Adhikari and Yin, 2020; Allel et al., 2020; An et al., 2020; Gupta and Gharehgozli, 2020; Pan et al., 2020; Djordjevic et al., 2021b; Hradsky and Komarek, 2021; Lorenzo et al., 2021; Markovic et al., 2021; Perone, 2021; Rontos et al., 2021; Salom et al., 2021; Singh et al., 2021; Wang et al., 2022)), many methodological obstacles complicate this type of research and often lead to conflicting conclusions of otherwise similar studies.

One obvious problem lies in often significant correlations between potentially relevant demographic predictors, making it challenging to disentangle their influences. This is further complicated by interactions between the variables and the nonlinear ways some of these predictors may influence COVID-19 observables. To distinguish between such delicate effects requires careful numerical analysis and sufficiently large COVID-19 data. State-of-the-art statistical and machine-learning methods can be effective if they are provided with sufficiently large, high-quality data. To train accurate models, one should collect relevant data from a large number of smaller regions. However, in the COVID-19 context, this comes with a trade-off: diverse regions tend to have inconsistent testing/reporting policies, and the data is often less reliable (or entirely unavailable) for smaller regions. In general, this dependence of COVID-19 observables (e.g., case counts and fatalities) on local policies (mostly on testing protocols and rules on which deaths are attributed to COVID-19) poses a problem in how to compare the data from various regions meaningfully. Even when policies reasonably coincide, making equal-time comparisons rarely makes sense since different counties of states belong to different phases of the epidemics curve.

Another methodological issue is to define the response variable, i.e., to quantify the precise aspect of the pandemic that we want to investigate and the appropriate proxy variables. In particular, there are two main, substantially different aspects of assessing the pandemic effects: i) Analyzing virus transmissibility, i.e., how rapidly it spreads in the community, which is necessary to understand the evolution of COVID-19 case numbers, and ii) Investigating SARS-CoV-2 severity - i.e., understanding individual hospitalization/morbidity/mortality risks, what causes differences in infection severity, and identifying subpopulations or regions more prone to severe forms of the disease. Of the two, much more effort has been devoted to the former, while the studies investigating the disease severity face the additional problem of choosing a relevant severity measure intrinsically independent of the transmissibility. For example, using COVID-19 fatalities is not suitable, despite being often used in this context (Wu et al., 2020; Moreira et al., 2021) since it is strongly correlated with COVID-19 prevalence (i.e., transmissibility) in the population (Markovic et al., 2021) - qualitatively, a larger number of cases (larger transmissibility) also leads to a larger number of fatalities.

In this paper, we focus on the problem of COVID-19 severity to address the above caveats. As the dataset, we collect COVID-19 time series (of case numbers and deaths) with values of over one hundred diverse sociodemographic variables for more than 3000 U.S. counties. This dataset has optimal properties in the sense of being both large and reasonably uniform (in the sense of COVID-19 policies). That is, all considered regions belong to the same country (and therefore have reasonably uniform policies). To focus on the influence of sociodemographic and economic factors, i.e., to neglect the complex influences of vaccination and different virus strains, we concentrate on the first epidemic wave – though, in the future, our study could also be extended by including suitable predictors for these factors. For each county, we estimate a well-defined measure of severity alone, which is a priori independent of the virus transmissibility. This measure, denoted as m/r, was introduced in Markovic et al. (2021) and is based on epidemiological modeling, representing the ratio of population-averaged mortality to recovery rates. Intuitively, the faster rate of dying from COVID-19, and slower recovery rate, relate to larger severity.

We apply several machine learning techniques to identify which demographic variables are relevant predictors of the m/r severity measure. In particular, we use repetitive rounds of (relaxed) Lasso and Elastic net linear regressions (with feature selection and regularization) and Random Forest and XGBoost, implementing ensembles of weak learners (decision trees). Random Forest and XGBoost can also accommodate highly nonlinear relations of the response to predictors and their interactions. Both can assign importance to the predictors, allowing for straightforwardly selecting significant predictors (with all other advantages of these techniques). Finally, we will also use a recently popularized (within the Uber platform (Zhao et al., 2019)) mRMR (minimal Redundancy Maximal Relevance) feature selection method, allowing better dealing with correlated datasets. mRMR will be integrated into Random Forest and XGBoost, which combines the advantages of these methods with mRMR. Overall, we carefully devise several state-of-the-art feature selection methods, intending to start from a large number of sociodemographic factors and, in an unbiased way (without prior assumptions), determine the most important predictors directly from the data. While machine learning has been successfully applied to a number of COVID-19-related problems, such as disease diagnosis and prognosis (Alizadehsani et al., 2021; Mahdavi et al., 2021; Amini et al., 2022; Kamalov et al., 2022; Rajab et al., 2022; Ramírez-del Real et al., 2022; Yousefzadeh et al., 2022) it was to our knowledge less frequently applied in the ecological study design (a transverse comparison of geographical regions) as done here (Wang et al., 2021).

The analysis presented here is also helpful from another perspective. In Markovic et al. (2021), we studied COVID-19 severity based on U.S. states instead of counties. Comparing the results of these studies can provide an important insight into the possible effects of spatial resolution (from 51 states to over 3000 counties) on the obtained results. To our best knowledge, it is currently unresolved what happens with conclusions of ecological regressions (transverse/crossectional study design across different regions employed here) in a transition from a smaller number of spatially larger geographic regions to a substantially larger number of smaller regions. Consequently, our study can also aid a better understanding of the implications of ecological regression study design, particularly in the context of machine learning applications.

## 2 Materials and Methods

### 2.1 Data collection

Demographic data at the county level were collected from several sources. The demographic composition of the U.S. population by gender, race, and population under 18 and over 65 was taken from the U.S. Census Bureau website integrating multiple different reports (U.S. Census Bureau, 2020). Information about population behavioral health risks at the county level was taken from the County Health Rankings website (County Health Rankings, 2020). The number of hospital beds and emergency unit capacity per county was obtained from the Homeland Infrastructure Foundation-Level Data (HIFLD) website (Homeland Infrastructure Foundation-Level Data, 2020). Poverty, deep poverty, median household income, per capita income, number of households, and predictor variables describing various levels of education on the county level were downloaded from the U.S. Department of Agriculture website resource Atlas of Rural and Small-Town America (U.S. Department Of Agriculture, 2021). Medical parameters such as hypertension, cardiovascular disease mortality, diabetes, obesity, inactivity, lower respiratory disease mortality, and daily smoking prevalence were downloaded from the Global Health Data Exchange website (Global Health Data Exchange, 2021). Individual county areas (U.S. Census Bureau, 2018) and exact FIPS codes (U.S. Census Bureau, 2011) were also downloaded from the U.S. Census Bureau website. Python scripts were used to map multiple county information sources using FIPS code values, and the resulting dataset is provided in Supplement Tables 1 and 2.

### 2.2 County severity measure calculation

Information about cumulative daily COVID-19 deaths and cumulative registered infection cases at the county level was retrieved from (Dong et al., 2020). From these case counts, the COVID-19 severity measure *m/r* was calculated as previously derived (Markovic et al., 2021) using our SPEIRD infection dynamics model (Djordjevic et al., 2021a, 2021b):

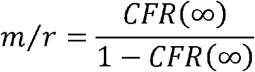

Here *CFR(*∞) is the Case Fatality Rate in saturation, i.e., calculated at the end of the epidemic wave. *CFR* corresponds to the ratio of cumulative fatalities and case counts, where both quantities are calculated at the end of the wave. To estimate its saturation value, without relying on a single date for the wave end, we use a mean CFR value for the time interval at the end of the wave when the case counts (and correspondingly also CFR) enter saturation. This time interval (end of the wave) was estimated at the level of states and then associated with the corresponding counties (see Supplement Table 3), as it was shown that the wave intervals could be inferred more accurately from larger (conglomerated) spatial units (Vilar and Saiz, 2021). For more details on the derivation of m/r, see Supplementary methods.

In Markovic et al. (2021) it was shown that the m/r measure is independent of transmissibility, which is also evident from the direct (though nonlinear) relationship between m/r and CFR (due to the fact that CFR is, per se, independent from the frequency of the virus transmission). Therefore, our measure does not depend on the rate at which the epidemic spreads, and is consequently independent of the social distancing measures and/or quarantine. It also does not depend on the epidemic phase since it is a function of CFR at the end of the epidemic wave (when both the number of fatalities and cumulative case counts have stabilized). Additionally, the m/r value is not expected to significantly depend on the testing policies. That is, while both the cumulative number of (detected) COVID-19 deaths and the cumulative (detected) case counts depend on the volume of testing, their dependence is qualitatively of the same manner: fewer tests will result in lower case counts but also in more COVID-19 deaths that failed to be attributed to the pandemic. Thus, these two effects tend to cancel each other.

Several other severity/fatality measures have been proposed so far, including the total number of fatalities, as the simplest to obtain, yet inadequate measure, which is highly correlated with the total number of detected cases, making it impossible to distinguish the severity from the transmissibility of the disease (Markovic et al., 2021). Some other, more promising approaches found in the literature include the use of CFR and its variations, such as delay-adjusted CFR (Yeoh et al., 2021). These measures, however, do not have a clear mechanistic interpretation (Böttcher et al., 2020), as they are not derived from a dynamic/mechanistic model of the disease spread.

Since CFR at the end of the first peak for COVID-19 has a relatively small value for most counties (∼10^−2^), from the equation above follows that in such cases m/r and CFR(∞) have similar values, so m/r in principle leads to the robust results compared to other measures (but only provided that CFR for these measures is calculated in saturation, i.e., at the end of the peak). However, this does not have to be the case for other infectious disease with potentially higher CFR, for which the difference between m/r and CFR(∞) would be more drastic, particularly since m/r is a nonlinear function of CFR(∞). In such a case, and for the reasons stated above, the use of m/r as the severity measure is more adequate. Even in the case of low CFR, and as explained above, using m/r has the following advantages: i) In distinction to ordinary CFR, CFR in m/r expression is calculated at saturation (end of the peak), which naturally follows from m/r derivation and makes the measure independent of the epidemic phase. ii) The measure has a clear mechanistic interpretation and is inherently independent of transmissibility (and by that, also of the effects of epidemiological policies and interventions), which further simplifies the result interpretation. Based on that, using m/r as the response variable in ecological regressions applied to epidemiological problems is preferable. In addition to the study of COVID-19 severity determinants at the level of USA states in Markovic et al. (2021), the measure was also successfully applied at a global level to better understand the apparently puzzling relationship between Global Health Security Index (GHSI) and COVID-19 mortality in different world countries (Markovic et al., 2022).

### 2.3 Data processing

All assembled variables were subjected to standard transformations of different strengths and directions (square, square root, cubic root, logarithm, negative square root, negative cubic root, and negative logarithm) to reduce the skewness of the data and bring them closer to normal distribution. For each variable, the transformation that minimizes the absolute value of the skewness from the Python SciPy library (Virtanen et al., 2020) was chosen, which was automated by a custom Python script. The county severity measure was transformed using the square root function (also chosen to minimize skewness).

Outliers were identified as being outside three median absolute deviations (MADs). After applying transformations, outliers were substituted with the corresponding variable median values. Transformations and subsequent outlier substitution by median values removed heavy distribution tails (observed for some variables) so that the distributions were brought closer to normal.

### 2.4 Model hyperparameter tuning

Processed data was split into training and validation sets (80-20). The validation set was set aside, while the training set was used for hyperparameter selection (through 10-fold cross-validations) and final model training. To select optimal hyperparameter values, we put them on an extensive grid (specified below for each model) and chose the parameter combination leading to the smallest cross-validation MSE (Mean Squared Error). Alternatively, to obtain sparse models (see below), hyperparameter combinations within one standard error of minimal MSE were considered. The data were standardized (the mean subtracted and divided by the standard deviation) in each cross-validation round. The hyperparameter grid search results are provided in Supplement Tables 4-13. Final models were trained on the entire training dataset with the previously selected optimal hyperparameter values. MSE calculated on the validation set was compared to (approximately) agree with the training set MSE as a consistency check.

### 2.5 Lasso regression

Lasso regression applies L1 regularization (Hastie et al., 2009), controlled by the λ hyperparameter value. Hyperparameter grid search was performed as described in 2.4., with exponential grid spacing and maximal λ value corresponding to all zero coefficients. λ values that lead to minimal cross-validation MSE within one standard error were selected. λ for training the model on the entire training dataset corresponds to the maximally sparse Lasso model (i.e., largest λ) within these values. Non-zero coefficients were extracted from the model.

### 2.6 Elastic net regression

Elastic net regression applies L1 and L2 regularization (Zou and Hastie, 2005; Hastie et al., 2009), which are controlled by λ and α hyperparameter values. ElasticNet Scikit-Learn library (Pedregosa et al., 2011) was used. α parameter was put on a linear grid in the range (0,1), and for each α parameter, the range of λ values was selected as described in 2.5. This resulted in a 2-dimensional grid, searched as described in 2.4. Hyperparameter combinations within one standard error of cross-validation MSE were selected. Among these, (λ, α) combination that leads to a maximally sparse Elastic net model was chosen to train the model on the entire training dataset.

### 2.7 Random Forest regression

For Random Forest regression, minimal leaf size and maximal tree depth were used as hyperparameters (Breiman, 2001; Hastie et al., 2009). The number of regression tree estimators was set to 600. Hyperparameter grid values that correspond, respectively, to minimal leaf size and maximal tree depth are: {1, 2, 4, 8, 16, 32, 64, 128, 256, 512, 1024}, {1, 106, 211, 316, 421, 526, 631, 736, 841, 946, 1051, 1156, 1261, 1366, 1471, 1576, 1681, 1786, 1891}. A 2-dimensional grid was constructed, and a search was performed as described in subsection 2.4. A hyperparameter combination that corresponds to minimal test MSE was selected. As several combinations correspond to the minimum, the one with the smallest maximal tree depth (corresponding to the shallowest tree) was selected. The Random Forest model was then trained on the whole training set, and predictors with greater than mean feature importance were selected.

### 2.8 XGBoost regression

XGBoost regression model learning rate, maximal tree depth, and the number of tree estimators were tuned hyperparameters (Friedman, 2001; Hastie et al., 2009; Chen and Guestrin, 2016). Hyperparameter values in the previously defined order are: {0.5, 0.1, 0.15, 0.2, 0.25, 0.35, 0.5}, {1, 2, 3, 4, 5, 6, 7, 8, 16, 32, 64, 128, 256, 512}, {15, 30, 45, 60, 75, 90, 100, 115, 130, 145, 160, 175}. A 3-dimensional grid was constructed, and a search was performed as described in 2.4. A Hyperparameter combination corresponding to minimal test MSE was selected and used to train the XGBoost model on the entire training dataset. Predictor variables with greater than mean feature importance were selected.

### 2.9 Relaxed models

Relaxed models (Hastie et al., 2009) were implemented through a two-step iterative training process. The first training step is described in the sections above (Lasso, Elastic net, Random Forest, and XGBoost regression). Input data for the first step contains an entire dataset with all 115 predictor variables. Hyperparameter values are optimized after the first round as described above, and predictor variables are selected for each model based on non-zero coefficients (Lasso and Elastic net) or greater than mean feature importance for Random Forest and XGBoost. In the second iteration, the input dataset contains only predictor features selected by the first iteration, and model training is repeated in the same way as for the first iteration. Second iteration (relaxed) models are further used to extract the final predictor importance and coefficients. In the Supplement Figures, we provide importance estimates for all predictors.

### 2.10 Minimum redundancy maximum relevance predictor selection

Minimum redundancy maximum relevance (mRMR) is an algorithm for selecting the minimal-optimal subset of predictor variables (Ding and Peng, 2005; Zhao et al., 2019). In mRMR implementation (Mazzanti, 2022), F-statistics was used to assess association with the response (relevance) and mean Pearson correlation between predictors to assess redundancy. mRMR regression returns top *n* selected features, where *n* was added as an additional hyperparameter to Random Forest and XGBoost regressions, with the grid values: {5, 10, 15, 20, 25, 30, 40, 50, 65, 80, 100, 115}. For each grid value, (1 – minimal MSE) was plotted, and the number of features was selected when the plot approached saturation. This number of features and other hyperparameter values corresponding to minimal cross-validation MSE were used to train Random Forest or XGBoost models on the entire training dataset. Predictor variables with greater than mean feature importance were selected.

## 3 Results

We started by assembling an extensive set of sociodemographic and medical variables for USA counties (115). The entire dataset is provided in Supplement Tables 1-3. The main challenge for the data analysis is a large number of input variables from which we should select the most important predictors of disease severity. While this allows for an unbiased selection of factors that can contribute to the disease severity at the level of counties, a large majority of the initial set of variables likely do not significantly contribute to the response. Therefore, keeping them in the analysis may lead to a large noise and, consequently, model overfitting. On the other hand, several sociodemographic factors can genuinely contribute to explaining severity, so multivariate analysis, in which one controls for simultaneous effects of these variables, is necessary. Consequently, we start with linear regression methods with regularizations and variable selection, Lasso (Hastie et al., 2009) and Elastic net (Zou and Hastie, 2005; Hastie et al., 2009). Both methods can exclude redundant variables that do not significantly contribute to m/r. To reduce the effect of noise, both algorithms were implemented in the so-called relaxed procedure (Hastie et al., 2009), consisting of two iterations. In the first iteration, the algorithm is trained on all predictors. A hyperparameter combination within one standard error of cross-validation MSE that led to a maximally sparse model was chosen. Only non-zero coefficient predictors are used in the second training iteration to reduce noise influence on the model. Taken together, the variable selection implemented through Lasso and Elastic Net, together with the relaxed model selection procedure, allowed reducing multicollinearity by removing redundant variables.

Results of the Lasso and Elastic net regressions are presented in Figure 1. Hyperparameters in both models are optimized on the grid through cross-validation so that the resulting model corresponds to maximal prediction accuracies on new datasets. Note that, as the data was standardized before the regression, the obtained regression coefficients can be interpreted as the importance of the given feature in explaining COVID-19 severity, while the coefficient’s sign indicates the influence’s direction. Both methods lead to similar results. Population density is singled out as the severity predictor with the highest importance, followed by the percentage of Black females. Both predictors positively affect m/r, i.e., higher population density and Black female percentage are related to higher disease severity. Of the predictors with somewhat lower importance, traffic volume is negatively associated with the disease severity, while PM air pollution and high housing costs (an indication of poor socioeconomic conditions) are positively associated with the severity. However, the importance of these three features is notably smaller than the importance of the population density and percentage of African Americans. Note that “Black female” and “Black male” variables are highly correlated (Pearson Correlation Coefficient of 0.93), which in practice makes them hardly distinguishable and redundant. Due to this, in the text we merge/consolidate them as a measure of African American population prevalence (African Americans).

**Figure 1.**
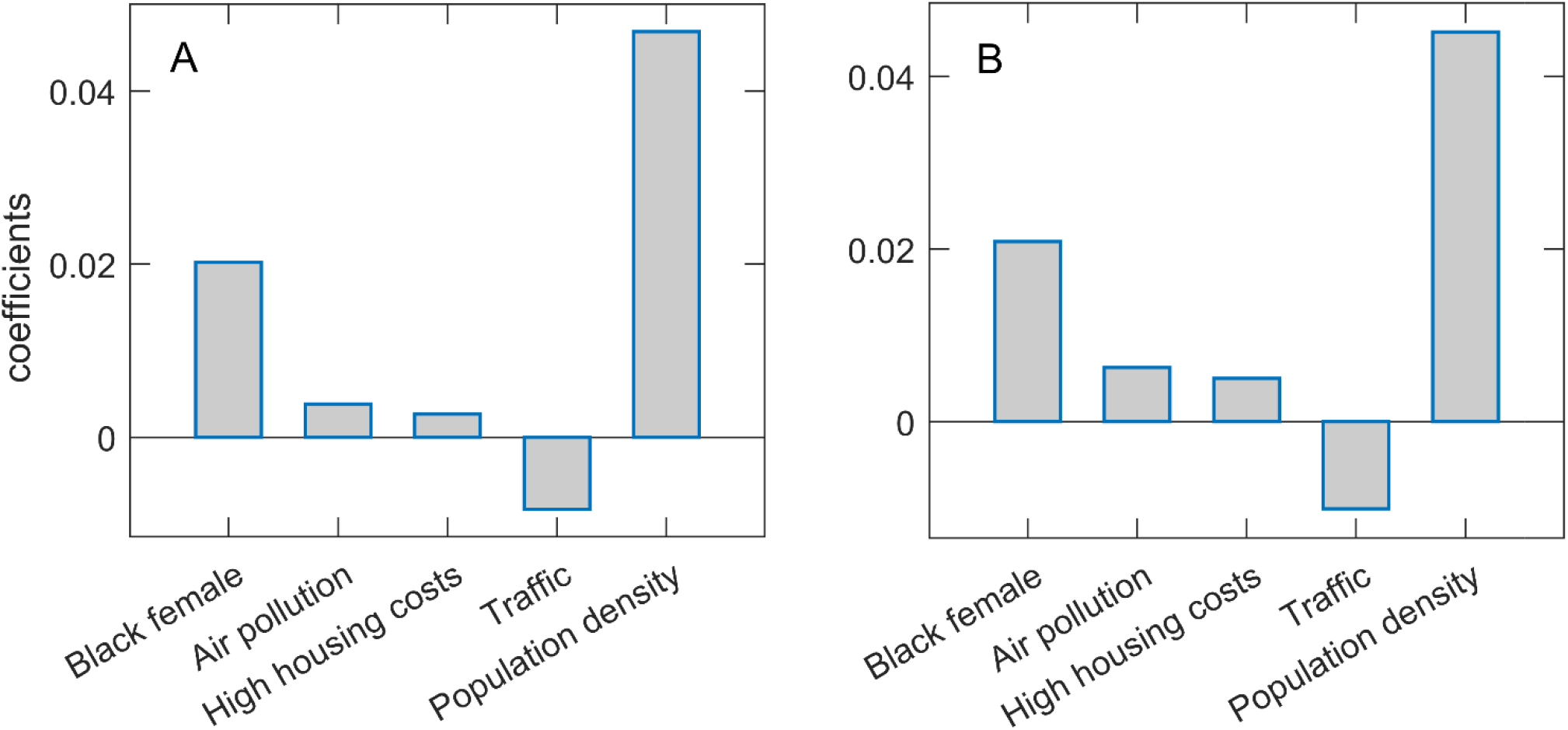
Predictor selection by Lasso and Elastic net. The regression coefficients, being a measure of the variable importance in explaining m/r, are shown for **(A)** Lasso and **(B)** Elastic net. The selected variables are indicated on the x-axis, while the y-axis corresponds to the coefficient’s values.

Lasso and Elastic net correspond to linear regression analysis. However, in reality, the predictors may have a highly nonlinear relationship with the output, while interactions between different predictors in the model may also occur. Linear regressions cannot account for such effects. Thus, we next used the ensembles of weak learners (decision trees), i.e., XGBoost and Random Forest. Another advantage of these methods is that they can better handle multicollinearity, particularly when redundant variables are removed (i.e., the most relevant variables selected), before training the ensembles of the decision trees. We extensively optimized (cross-validated) both methods over a large hyperparameter grid. We again employ both methods in the relaxed setup to reduce noise influence, i.e., only the predictors with importance above the mean (standardly used threshold) in the first round are used as the input in the second round.

Figure 2 presents feature importance in Relaxed Random Forest and XGBoost. Again, robust results consistent across the two methods were obtained, where by far the highest relative importance is assigned to population density, followed by the Black female variable. These results are consistent with those previously obtained by Lasso and Elastic net regressions. Besides these two features, which are clearly above the importance threshold in both methods, traffic volume and high housing costs appear with values barely above the threshold in XGBoost.

**Figure 2.**
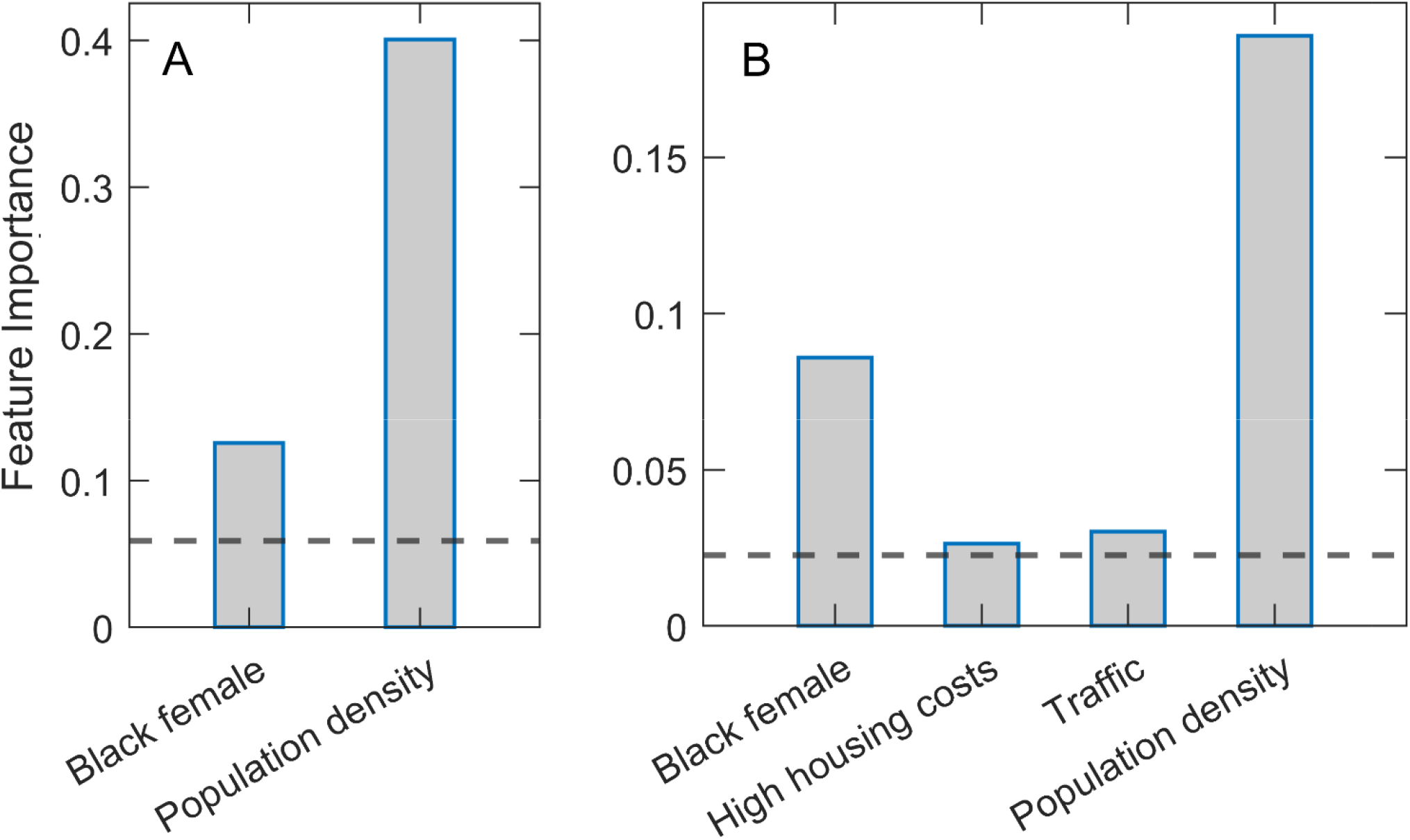
Estimated variable importance in relaxed Random Forest and XGBoost methods. **(A)** Random Forest and **(B)** XGBoost methods are implemented in the relaxed procedure, where only variables above the importance threshold in the second round are shown. The estimated variable importance is shown on the vertical axis. The horizontal line indicates the standard threshold for the significant predictors (corresponding to the predictor mean).

Mutual correlations between the predictors in the dataset are another complication. To address this, we integrate the mRMR method into Random Forest and XGBoost methods. The method was initially introduced by Ding and Peng (2005) but recently gained popularity with its implementation within the Uber machine learning platform (Zhao et al., 2019). In essence, mRMR ranks the variables to how well they are associated with the response and how much they are redundant (where high correlations with other predictors decrease the predictor rank). In the Uber platform, the method was integrated only in Random Forest, and fixed (preselected) hyperparameter values were used, likely to reduce computational time in a time-sensitive setup. Instead, we here carefully optimize hyperparameters by cross-validation on an extensive grid. The number of selected predictors in this cross-validation is also treated as a hyperparameter (see Methods). We also implement mRMR within XGBoost, in addition to being implemented in Random Forest.

Results of Random Forest and XGBoost with integrated mRMR methods for variable preselection are shown in Figure 3. Optimal selection of the number of variables was made through the plots on the left-hand side of the figure panel (3A and C), where the prediction accuracy (assessed on the testing set in cross-validation) is shown vs. the number of selected variables. Above a certain number of included variables, the prediction accuracy enters saturation, which we use for selecting the number of variables for training the final model. The number of retained features was 25 for Random Forest and 38 for XGBoost. Figures 3B and D (the right side of the panel) again show the dominant importance of Population density and the Black female variable. While in Random Forest, we obtain no other features above the importance threshold, several features in XGBoost have importance estimates above the mean importance value. Most notably, the percentage of the rural population, high housing costs, percentage of white males, and traffic volume. We will see that most of these variables significantly correlate with the two main predictors.

**Figure 3.**
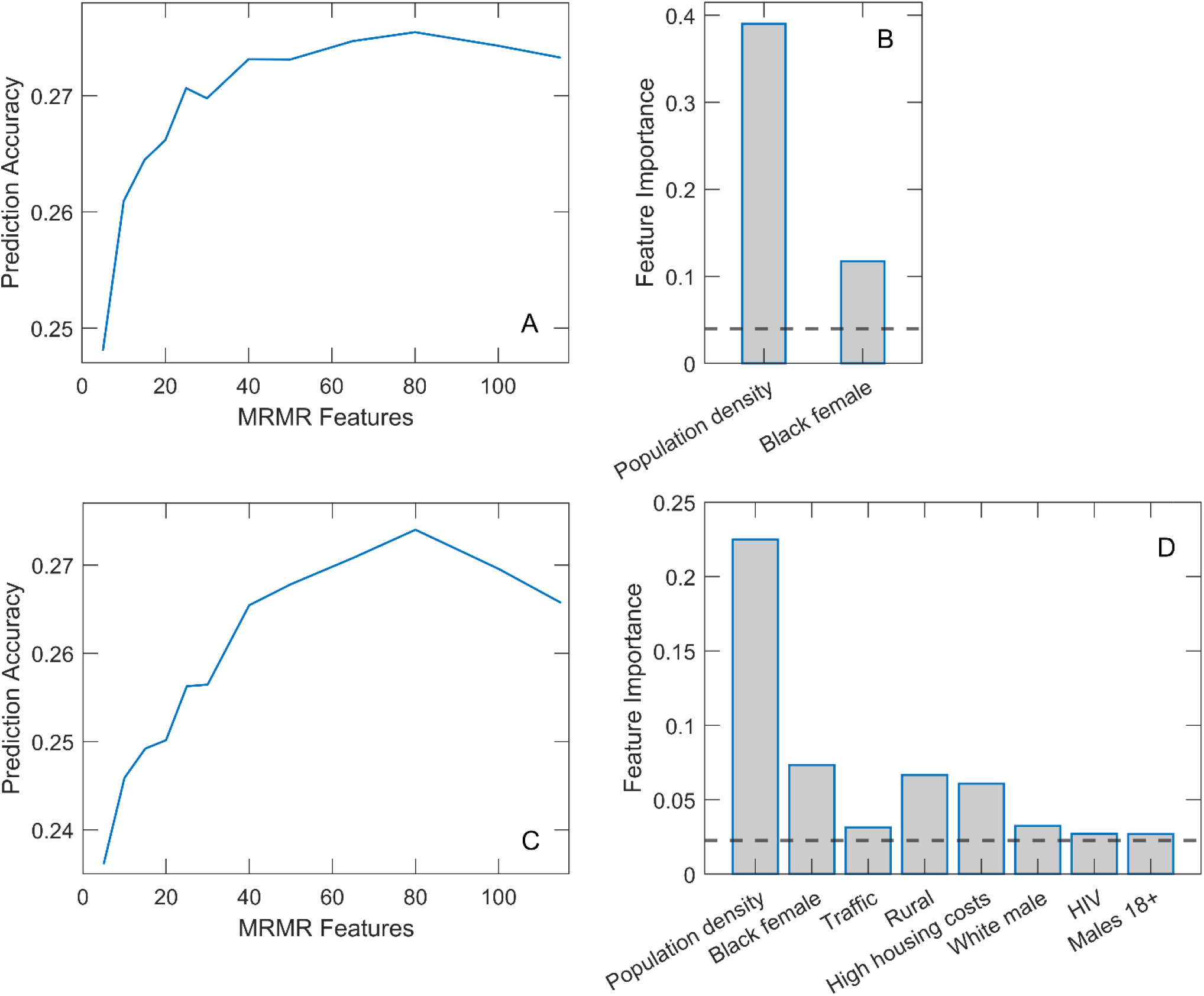
Relaxed Random Forest and XGBoost with mRMR feature selection. Feature selection for Random Forest **(A)** and XGBoost **(C)** by mRMR method. Feature importance estimates in Relaxed Random Forest **(B)** and XGBoost **(D)**, where only variables above the importance threshold in the second round are shown.

Interestingly, only two predictors (Population density and Black female) were robustly singled out from 115 variables used in the initial input in the analysis. We finally assess the correlation of these two variables with the other variables to discuss factors related to the two main predictors associated with m/r. The variables with the highest values of the correlation coefficients are shown on the bar plots in Fig. 4. All these variables have a statistically highly significant correlation (P∼10^−100^). These correlations are further discussed in the next section.

**Figure 4.**
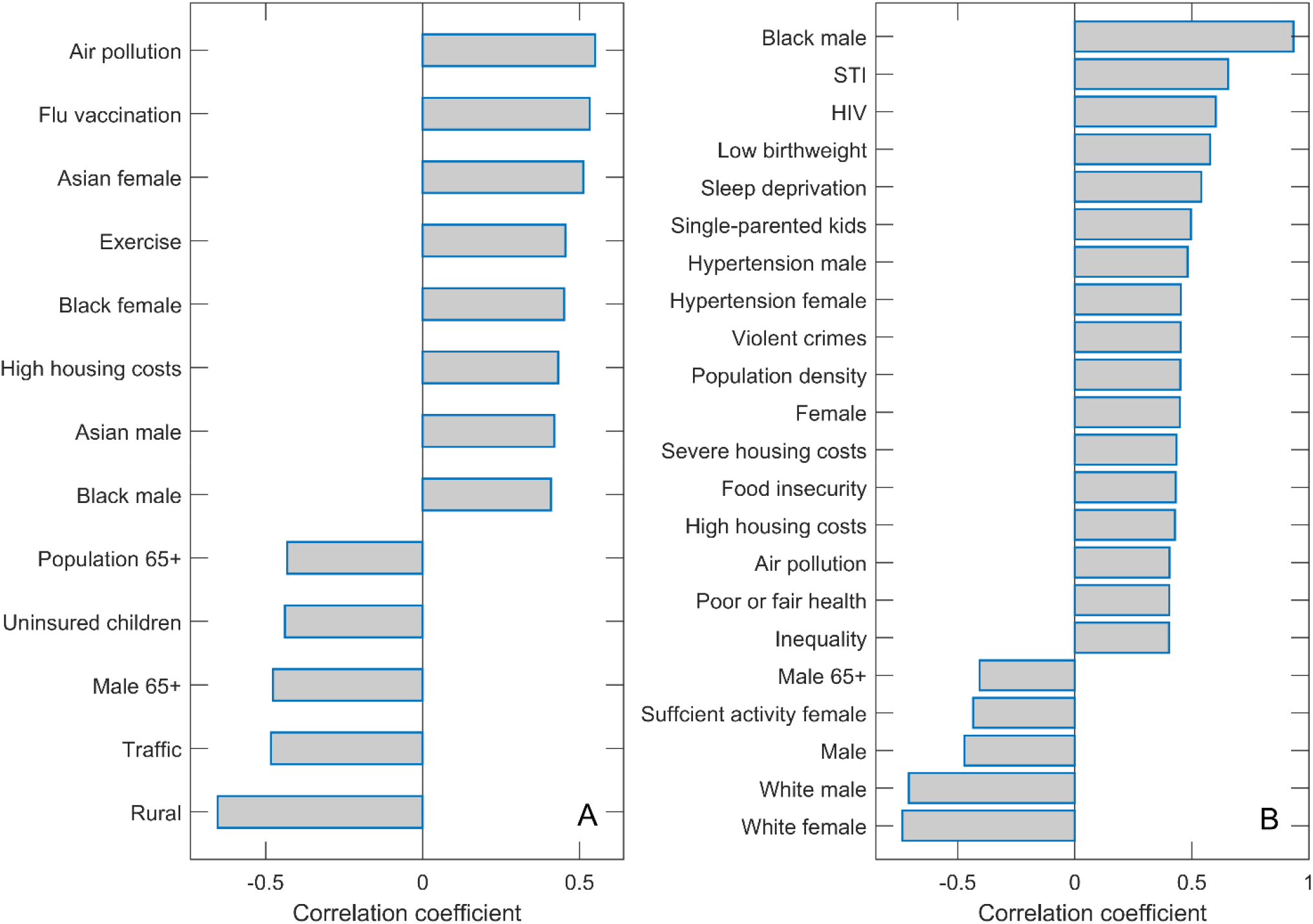
Correlation of the population density and Black female with the other variables. Names of the variables with magnitudes of Pearson correlations (either positive or negative) larger than 0.4 are shown on the bar plots for **(A)** Population density and **(B)** Black female.

## 4 Discussion

In our large-scale high-resolution study (county-level with many predictors), we robustly obtain population density and percentage of Black females as the COVID-19 severity predictors with the highest importance in regressions. For discussion, we correlated these variables with the other predictors and selected those with the highest correlations.

This can be informative when trying to understand our somewhat surprising result: only two variables were clearly selected among a large number of starting predictors. By considering these correlations, we may also better understand possible factors that contribute to these two variables being clearly distinct in their association with COVID-19 severity.

As the Black female variable is strongly positively correlated with the Black male variable, it can be considered as a measure of the percentage of African Americans of both genders. Furthermore, the fraction of the Black population is strongly negatively correlated with the proportion of the non-Hispanic white population and positively correlated with the Asian American and Hispanic populations. The Black female variable can be, thus, considered a signature of the minority population, which we found strongly positively associated with COVID-19 severity. Indeed, this association also holds for the Hispanic population, who, despite having (on average) higher life expectancy (County Health Rankings, 2020) compared to non-Hispanic whites, suffered the highest drop in life expectancy due to COVID-19 compared to any other ethnicity (Woolf et al., 2021).

The positive association between the percentage of African Americans and the severity of COVID-19 has already been documented (Azar et al., 2020; Thebault et al., 2020) and discussed in the context of several health and social factors. These are the same factors that show up in our analysis through the correlations of Blacks with other variables. First, Blacks are strongly correlated with several determinants of poverty and disadvantaged population, such as the prevalence of sexually transmitted infections (STIs), violent crimes, different housing problems, and smaller homeownership. COVID-19 severity has also been associated with determinants of the disadvantaged population outside the USA (Gao et al., 2022). Secondly, they are strongly correlated with a number of medical factors, such as low birth weight, insufficient sleep, hypertension, cardiovascular diseases, and generally poor health. These medical conditions are well-known COVID-19 risk factors, as extensively discussed in the literature (Ssentongo et al., 2020; Ahmadi et al., 2021; Crispi et al., 2021; Du et al., 2021; Saleh et al., 2022; Zhang et al., 2022). We next focus on sociodemographic factors, whose interpretation may be less evident.

Although the direct association between the prevalence of violent crimes and COVID-19 severity is unlikely, this variable can be interpreted as another measure of socioeconomic deprivation, as it is established that both poverty and income inequality are positively associated with the rate of violent crimes (Hsieh and Pugh, 1993; Kennedy et al., 1998). While a higher rate of violent crimes is correlated with a larger proportion of African Americans, violent crimes are more likely to be class-related (Smith et al., 2021) than associated with African Americans per se. Violent crime areas may also lead to high-stress levels (Berman et al., 1996; Ellen et al., 2001; County Health Rankings, 2020), which can damage health and be the underlying cause of a series of chronic conditions, such as hypertension (Zimmerman and Frohlich, 1990; Ellen et al., 2001) or obesity (Conklin et al., 2019), which are both well-known risk factors for the severe outcome of COVID-19 (Kwok et al., 2020; Du et al., 2021).

Housing issues, such as severe or high housing costs and a low homeownership percentage, indicate poor socioeconomic conditions (Dunn, 2002; County Health Rankings, 2020). Race differences also play a role in homeownership, as it is much lower among African Americans than non-Hispanic Whites (Jackman and Jackman, 1980). Households affected by housing issues would probably lack access to healthcare, as they may be unable to pay for it (Carroll et al., 2017). In the pandemic context, members of such households might not receive proper medical care, fail to timely seek medical attention, or be unable to afford the appropriate treatment and medications. Prevalence of STIs, defined as the number of newly diagnosed Chlamydia cases per 100,000 population, is also correlated with the Black female variable, which is not surprising, as it has been shown that African American adolescent women are disproportionately affected by Chlamydia (Cooksey et al., 2010). The prevalence of Chlamydia and other STIs can thus be viewed in the context of health inequality (County Health Rankings, 2020).

Therefore, all sociodemographic variables significantly correlated with Blacks correspond to underserved communities. This suggests that Black female was singled out by our regressions, not as a single severity predictor but as the variable that best captures most of these effects, indicating that minorities and socially disadvantaged populations were disproportionately severely affected by COVID-19, which is coherent with the results of several other studies (Dyer, 2020; Tirupathi et al., 2020; Arasteh, 2021; Chen and Krieger, 2021; Tai et al., 2021). Additionally, middle-aged Black females have already been recognized as the group with the highest disease burden in Mississippi (Martin and Garrett, 2022). This could be related to the higher prevalence of obesity in this social group (Martin and Garrett, 2022) or a relatively high percentage of Black females who are essential workers (Sugg et al., 2021) working in an environment with a probability of high viral exposure. High initial viral inoculum at the workplace could also lead to higher disease severity (Burgess et al., 2020). Since people in disadvantaged areas are more likely to be “essential workers” working in environments with a high risk of COVID-19 exposure while simultaneously having limited access to healthcare (Oronce et al., 2020), the obtained associations with the disease case counts are not surprising. However, as our severity measure is independent of transmissibility, our result is not a mere consequence of a larger COVID-19 exposure but rather a consequence of the interplay of medical and sociodemographic factors discussed above.

Population density appeared as, by far, the most significant predictor of COVID-19 severity. The variable with the highest correlation with population density is air pollution. This likely points to an important factor behind the strong association of population density with disease severity. Namely, the link between air pollution exposure and respiratory diseases, and COVID-19, in particular, is well established (Ogen, 2020; Wu et al., 2020; Pansini and Fornacca, 2021).

In addition to pollution, the population density is also significantly correlated with Blacks, where potential contributions of this variable to the severity are discussed above. Other variables correlated with the Black female also appear to correlate with population density (STI, housing problems, insufficient sleep). Interestingly, another racial-related factor (non-White/White residential segregation), which did not turn out to be highly correlated with Blacks, now appears significantly correlated with the population density. Regarding minorities, the Asian population of both genders is also significantly correlated with population density. The variable with the highest negative correlation with the population density is Rural, so the population density is a good proxy for urban and metropolitan areas.

Apart from the influences via pollution and African Americans (and related variables discussed above), likely, population density is also, *per se*, a prominent risk factor. A strong, nonlinear association between the epidemic’s size and population density has already been proposed (Kermack et al., 1927) and empirically confirmed (Li et al., 2018). As higher population density inevitably leads to a much higher number of infected individuals in densely populated areas, the number of patients requiring hospitalization is more likely to quickly exceed the healthcare capacity. This effect of overcrowding, in this case, is dominant compared to the disparities in healthcare in rural areas, where population density is low. Namely, even though people in rural areas often struggle with poverty, lack of health insurance, and shortages in health professionals (Probst et al., 2004), a lower probability of exposure to the virus leads to the generally lower severity of the disease in these areas, so that healthcare facilities cannot quickly become saturated. Another possible explanation for the lower severity in rural areas is underreporting of COVID-19 deaths in these areas (Souch and Cossman, 2020). Namely, it has been determined that excess mortality not attributed to COVID-19 was higher in counties with a lower percentage of insured individuals, fewer primary care physicians, and more at-home deaths (Stokes et al., 2021). As most of these characteristics apply to rural areas (Probst et al., 2004), the reported cases and deaths likely do not correspond well to the actual situation.

Finally, this work provides an opportunity to compare the results of this high-resolution (county-level) analysis with our previous study at the state level (Markovic et al., 2021). While, in addition to different geographic resolutions, the two studies also use different variables – a larger number of (different) predictors are used here – interesting comparisons can still be made. First, predictors related to population density, African Americans, pollution, and prevalence of chronic diseases were obtained in that study. Although all these variables were directly selected at the state level, in the present study, pollution and chronic diseases were also identified via association with the two directly selected predictors. Also, at the state level, African Americans were less robustly selected, i.e., only in the analysis that considers nonlinearities and interactions between the predictors, while in this study, it was robustly selected as a major predictor.

The largest difference between the two studies is the effect of the population age, which was selected as a significant predictor (with the expected positive influence on severity) in (Markovic et al., 2021) but did not emerge as significant in this study. A higher proportion of African Americans and population density are associated with a younger population. It appears that, at the county level, Blacks are a much stronger signal associated with a younger population, which appears to conceal the age effect on severity. That is, counties with older populations will also have a smaller Black fraction, so they do not appear with higher m/r. At the state level, the variations of Blacks are lower (so that Blacks come out only in a more complicated machine learning analysis), which allows the age effect to come out. On the other hand, age has been clinically recognized as an important COVID-19 severity risk factor. This, therefore, shows that the analysis at lower and higher spatial resolutions are complementary, i.e., the smaller spatial resolution is not necessarily more accurate/relevant. One reason is that decreasing the size of the regions where the analysis is done also decreases the number of case counts, thereby increasing fluctuations and, consequently, the noise in the model. This consequently argues that, at least for some significant predictors, larger spatial resolution may clearly promote their proper identification.

## 5 Study limitations

We finally discuss some limitations of our study. Most importantly, while we here assembled a vast number of COVID-19 predictors, some factors that are likely very important (but would be hard to quantify) are clearly missing. In particular, our dataset consists of “static” variables and does not include “dynamic” decisions and factors that emerge during the pandemic, such as decisions on how to treat patients, medical protocols to be applied, motivation/training of medical staff, etc. In other words, static capacities or beneficial general conditions to fight pandemics may not necessarily translate to optimal decisions (and willingness to implement them), as has been well recognized in the case of, e.g., Global Health Security Index (Haider et al., 2020; Stribling et al., 2020). How to systematically include/quantify such highly complex factors remains to be seen.

On the other hand, a significant advantage of our study is that the epidemic intervention decisions (social distancing, quarantine, etc.) that impact the disease spread (transmissibility) (Hayashi et al., 2022) do not influence our severity measure (Markovic et al., 2021). This is because our severity measure m/r is independent of transmissibility, which does not apply to measures commonly used to quantify COVID-19 severity/mortality (such as the number of fatalities). We feel this is a considerable advantage of our study, as the actual effect of introduced intervention measures is hard to quantify (Soltesz et al., 2020). Also, as discussed in the Materials and Methods section, m/r is neither expected to significantly depend on the testing policies, since the variations in the volume of testing affect both the numerator and the denominator of the CFR in the same direction. However, the strict independence of m/r on testing policies would require that the influence of the testing coverage on the case counts is exactly proportional to its effect on the number of fatalities, which need not be the case. Possible larger deviations in this sense might affect some of our conclusions.

Meteorological variables are also not included in this study. While they may impact transmissibility (Salom et al., 2021; Lin et al., 2022), they are unlikely to significantly impact the disease severity/mortality, as explicitly obtained for the state-level analysis (Markovic et al., 2021). Also, another potential limitation is that we inferred the end of the peak time from states (and then applied them to corresponding counties). It was previously shown that the peak time range could be inferred more accurately in spatially larger (conglomerated) regions (Vilar and Saiz, 2021), though we cannot exclude some counties with different peak timing. However, this should not significantly impact our results since in Markovic et al. (2021), we showed that m/r enters saturation (i.e., is nearly constant) for an extended time period, so the results should not be susceptible to exact dates for m/r inference. More generally, however, this issue corresponds to the conundrum of using smaller vs. larger geographic regions, which is generally understudied and should be better explored in the future.

Finally, as with other machine learning studies (limited to exploring data associations), the significant predictors we identified do not necessarily have to represent a causal relationship. In particular, our analysis has singled out two demographic factors - the percentage of Black females and population density - neither of which seems to have a direct medical impact on the prognosis of the disease. While this is clear for the population density, it is also less likely (though not entirely impossible) that Black females are genetically, per se, more predisposed to severe outcomes. Therefore, in an attempt to point out possible causal associations, we extensively discussed our results both in the context of previous studies and by analyzing the correlations of these two factors with the rest of the collected data. Thanks to the specific nature of the identified factors and the large overall number of variables included in the study, we believe that our interpretation of the obtained results indeed reveals some of the main drivers behind variations in the observed COVID-19 severity. Nevertheless, even if the significant predictors only partially reflect direct causal relations, they are still valuable risk assessment factors. Moreover, they may point to potential mechanistic relations that future studies should explore.

## 6 Conclusion and Outlook

We addressed the challenging problem of identifying some of the potential main drivers of COVID-19 severity from a large set of assembled sociodemographic factors. We showed that machine learning methods with feature selection are well suited for this task, producing robust results across different methods. The combination of mRMR and ensembles of decision trees (Random Forest and XGBoost) seems particularly promising for similar tasks in the future, as it can simultaneously handle large, correlated sets of predictors, their interactions, and nonlinear dependences. We propose that this methodology is useful whenever there is a measure of interest (response) defined over a diverse set of geographic regions, and significant predictors of this measure (e.g., demographic, economic, medical variables, or their combinations) should be selected among many variables that initially seem potentially relevant. In the study of COVID-19 and any other emerging infectious disease, identifying potential transmissibility and severity determinants (and the consequent understanding of the nature of their relation to the response variable) is a very challenging problem that requires taking into account many potential risk factors. The combination of the mRMR approach (which offers a very efficient way of variable preselection while eliminating all the redundant variables), and the nonparametric, supervised machine learning methods based on the ensembles of decision trees (which are capable of selecting important features while taking into account possible nonlinear relation between the features and the response variable), can be very promising in resolving this problem, as our study illustrates.

In summary, our final result is simple and suggests that densely populated areas with a high proportion of minorities and disadvantaged populations are the main COVID-19 severity risk factors. The result is here obtained for the USA, but it is arguably more general. That is, the likely causes behind such result are disadvantaged populations, environmental factors such as pollution, and a potentially high increase of cases in densely populated areas that available medical resources might not match. These factors remain to be carefully investigated and understood in the future.

## Data Availability

All data produced in the present study are available upon reasonable request to the authors.

## 7 Acknowledgments

We thank Dusan Zigic and Magdalena Djordjevic for their help in assembling part of the input data. This work was partially supported by the Ministry of Education, Science and Technological Development of the Republic of Serbia.

## 9 Author Contributions

MD conceived the research. The work was supervised by M.D. and I.S. Data acquisition and analysis by M.T. Figures and tables made by M.T. and S.M. A literature search by S.M. Manuscript written by M.T. and I.S., with the help of M.D. and S.M.

